# Early prognostication of COVID-19 to guide hospitalisation versus outpatient monitoring using a point-of-test risk prediction score

**DOI:** 10.1101/2020.10.19.20215426

**Authors:** Felix Chua, Rama Vancheeswaran, Adrian Draper, Tejal Vaghela, Matthew Knight, Rahul Mogal, Jaswinder Singh, Lisa G Spencer, Erica Thwaite, Harry Mitchell, Sam Calmonson, Noor Mahdi, Shershah Assadullah, Matthew Leung, Aisling O’Neill, Chayya Popat, Radhika Kumar, Thomas J Humphries, Rebecca Talbutt, Sarika Raghunath, Philip L Molyneaux, Miriam Schechter, Jeremy Lowe, Andrew Barlow

## Abstract

**Introduction:** Risk factors of adverse outcomes in COVID-19 are defined but stratification of mortality using non-laboratory measured scores, particularly at the time of pre-hospital SARS-CoV-2 testing, is lacking.

**Methods:** Multivariate regression with bootstrapping was used to identify independent mortality predictors in a derivation cohort of COVID-19 patients. Predictions were externally validated in a large random sample of the ISARIC cohort (N=14,231) and a smaller cohort from Aintree (N=290).

**Results:** 983 patients (median age 70, IQR 53-83; in-hospital mortality 29.9%) were recruited over an 11-week study period. Through sequential modelling, a 5-predictor score termed SOARS (<u>S</u>pO2, <u>O</u>besity, <u>A</u>ge, <u>R</u>espiratory rate, <u>S</u>troke history) was developed to correlate COVID-19 severity across low, moderate and high strata of mortality risk. The score discriminated well for in-hospital death, with area under the receiver operating characteristic values of 0.82, 0.80 and 0.74 in the derivation, Aintree and ISARIC validation cohorts respectively. Its predictive accuracy (calibration) in both external cohorts was consistently higher in patients with milder disease (SOARS 0-1), the same individuals who could be identified for safe outpatient monitoring. Prediction of a non-fatal outcome in this group was accompanied by high score sensitivity (99.2%) and negative predictive value (95.9%).

**Conclusion:** The SOARS score uses constitutive and readily assessed individual characteristics to predict the risk of COVID-19 death. Deployment of the score could potentially inform clinical triage in pre-admission settings where expedient and reliable decision-making is key. The resurgence of SARS-CoV-2 transmission provides an opportunity to further validate and update its performance.

## INTRODUCTION

Rapid and accurate prediction of the probability of adverse clinical outcomes is central to the management of global outbreaks of infection. [1-3] Stratification by predicted risk, most commonly for death, can support clinical judgement and potentially assist clinicians in community settings to decide how urgently to refer patients to hospital. Used appropriately, predictive scores can also help inform treatment-related decision making. The pandemic caused by severe acute respiratory syndrome coronavirus 2 (SARS-CoV-2) lends itself to predictive modelling by having a large at-risk population and a high adverse event rate including death.

Although the recent incidence of COVID-19 has decreased in some parts of the world, many countries are already experiencing a ‘second wave’ of new cases. It is widely anticipated that viral transmission will continue to surge in the months ahead, particularly with the onset of winter in the northern hemisphere. Not all patients infected with SARS-CoV-2 will require hospitalisation but even among those who initially experience mild symptoms, a sizeable proportion remain at risk of subsequent life-threatening clinical decline. The availability of a practical pre-hospital predictive tool to triage patients for safe discharge to an outpatient (virtual) monitoring system versus direct admission to hospital for observation or treatment would be highly advantageous.

Reliable prediction tools to differentiate between levels and sites of clinical care already exist and have been successfully implemented in pre-hospital practice. For example, both the CURB-65 and CRB-65 scoring systems for the assessment of community-acquired pneumonia include recommendations for out-of-hospital care. [4, 5] Recent research by the ISARIC-4C Consortium has provided an accurate tool to similarly prognosticate for COVID-19-attributed death in hospitalised patients but its reliance on laboratory-measured indices limits its applicability outside the institutional environment. [6] Prognostic evaluation of individuals with suspected SARS-CoV-2 infection at the time of diagnostic testing is potentially achievable but has yet to be examined.

Our objective was to develop and evaluate an easy-to-apply and accurate prediction score that could identify, early in the illness course, which patients infected with SARS-CoV-2 might benefit from an urgent hospital assessment or could be safely monitored in their own home. To develop the initial risk score, we used multivariate logistic regression to explore the relationships between a large panel of candidate predictors and COVID-19 death. Iterative modelling resulted in a pragmatic predictive score based on five widely available patient variables. The scoring of patients against these selected predictors permitted three distinct risk classes to be defined. The performance of the score was then assessed against two validation cohorts – a large and randomly selected subgroup of the ISARIC study patients and a smaller single-hospital cohort, the latter to better reflect local population characteristics and practice.

## METHODS

### Study design and characteristics of the derivation cohort

All individuals aged 18 or over who tested positive for SARS-CoV-2 nucleic acid by real-time reverse transcriptase polymerase chain reaction between 1^st^ March and 16^th^ May 2020 after presenting to the Emergency Department (ED) at Watford Hospital, West Hertfordshire NHS Hospitals Trust were prospectively recruited. Baseline clinical characteristics and investigation results were collected according to a pre-specified protocol. Patients were either referred to the virtual hospital (VH) for outpatient monitoring or admitted to a medical ward. The study was approved by the National Health Service Health Research Authority (20/HRA/2344; ethics reference: 283888).

### Laboratory, physiologic and radiographic data

All laboratory tests were performed as part of routine clinical care. Nasopharyngeal mucosal swabs for rRT-PCR were couriered to the regional UK Public Health England laboratory. Baseline vital observations included all the parameters recommended by the National Early Warning Score. [7] Chest radiographs acquired in ED were collated and scored at the end of the recruitment period.

### Location and level of care

Following presentation, patients who were clinically judged to have mild illness were referred to the virtual hospital for subsequent monitoring. To avoid missing early clinical deterioration in the post-assessment period, they were observed for up to 24 hours in hospital. Patients who remained admitted after the first 24 hours but who did not require additional respiratory support beyond wall-based oxygen were managed on designated medical wards. Where clinically indicated, continuous positive airway pressure (CPAP) was provided on such wards or on the intensive care unit (ICU); intubation and mechanical ventilation were undertaken on the ICU.

### Identifying predictors of death in the derivation cohort

The primary outcome of the study was hospital discharge or in-hospital death. TRIPOD recommendations were followed for multivariate model evaluation and reporting. [8] 75 baseline clinical and non-clinical variables were initially collected based on their reported association with COVID-19 and analysed by uni- and multivariate logistic regression with bootstrap resampling. [3-5,9] Candidate predictors of death were assessed for potential clustering effects and missing at random values were addressed by multiple imputation with chained equations (MICE), [9] with 10 – 20 random draws to account for data variability.

### Development and external validation of the clinical risk score

The large external cohort comprised a randomly selected sub-population of the ISARIC 4C derivation population (N=20,000 provided; 14,231 with complete data for scoring). The primary data of these individuals were submitted by 260 hospitals across England, Scotland and Wales to the prospective ISARIC World Health Organization Clinical Characterization Protocol UK (CCP-UK) study. [6] We also tested our score against a smaller population of SARS-CoV-2-positive cases from Aintree Hospital, Liverpool (N=303 provided; N=290 with complete data for scoring) as a single-setting validation control.

All performance metrics against external cohorts were analysed using the prediction score and not the regression model. In the initial stages, a preliminary score comprising 12 independent predictors of death, including Care Home residency, was developed; to enable external validation against the ISARIC cohort (which did not include residential data), Care Home status was excluded as a variable to yield an 11-predictor score. Its ability to discriminate for in-hospital mortality was assessed by the area under the receiver operating characteristic (AUROC). From this score, a condensed version comprising 5 clinical predictors was developed for pre-hospital application. Mortality cut-points at each risk level were assessed to define mild, moderate and high risk classes, followed by determination of positive and negative predictive values as well as sensitivity and specificity thresholds. Model performance was further assessed by calibration using a graphical representation of the Hosmer-Lemeshow ‘goodness-of-fit’ test to depict agreement between the expected (predicted) and observed (actual) outcome across the entire COVID-19 severity range in both validation cohorts. The summary relationship between the dependent variable (death) and different levels of disease severity in the external ISARIC population was expressed as McFadden’s *R*^*2*^.

### Statistical analysis

Categorical variables were expressed as frequency (%), with significance determined by the Chi-squared test. Continuous variables were expressed as median (interquartile range) or mean (standard deviation) and analysed by the t-test, Kruskal-Wallis or Mann-Whitney U test, as appropriate. Odds ratios (ORs) were assessed as unadjusted and adjusted values with respect to in-hospital death, the latter determined by multivariate regression with bootstrapping of 1000 resamples (10 Bootstrap). MICE was used to generate valid estimates of randomly missing values in the derivation model. A *P* value of <0.05 was considered statistically significant. All statistical analyses including risk modelling calculations were performed using STATA, version 16 (Stata Corp., Texas, USA).

## RESULTS

### Baseline demographic and clinical characteristics of the derivation cohort

983 patients (52.5% male) confirmed as SARS-CoV-2 rRT-PCR-positive were recruited over the 11-week study period. Five patients remained in hospital at the time of data censoring on 31^st^ May 2020. The median age of the cohort was 70 (IQR: 53 – 83; range 23 – 99); median age was lowest in the virtual hospital pathway (53; IQR 43 – 67) and highest amongst hospitalised patients who did not receive CPAP (77; IQR 61 – 86) (*P*<0.001) (appendix 1).

The commonest co-morbidities were hypertension (48.4%), pulmonary disease (30.0%), cardiac disease (26.6%), diabetes mellitus (23.6%), chronic kidney disease (CKD; 20.0%) and dementia (15.4%). Obesity, defined as BMI >30, was present in 24.7% (243) of the cohort, and associated with an unadjusted OR for death of 1.40 (95% CI: 1.18 – 2.68, *P* <0.05).

Overall, 294/983 (29.9%) patients died in hospital, the vast majority (97.3%) aged 50 or over. The mortality rates of different age brackets in the cohort (compared to the ISARIC and Aintree validation cohorts) are shown in appendix 2. The univariate OR for death increased with rising age, and was 14.86 (95% CI: 6.89 – 32.04) for those aged 70 – 79 and 20.87 (95% CI: 9.93 – 43.86) for those aged 80 or older (table 1). When stratified by maximal levels of care, mortality rate was lowest in the VH (1.8%) and highest in the ICU group (62.1%) (*P*<0.001) (appendix 3).

**Table 1.**
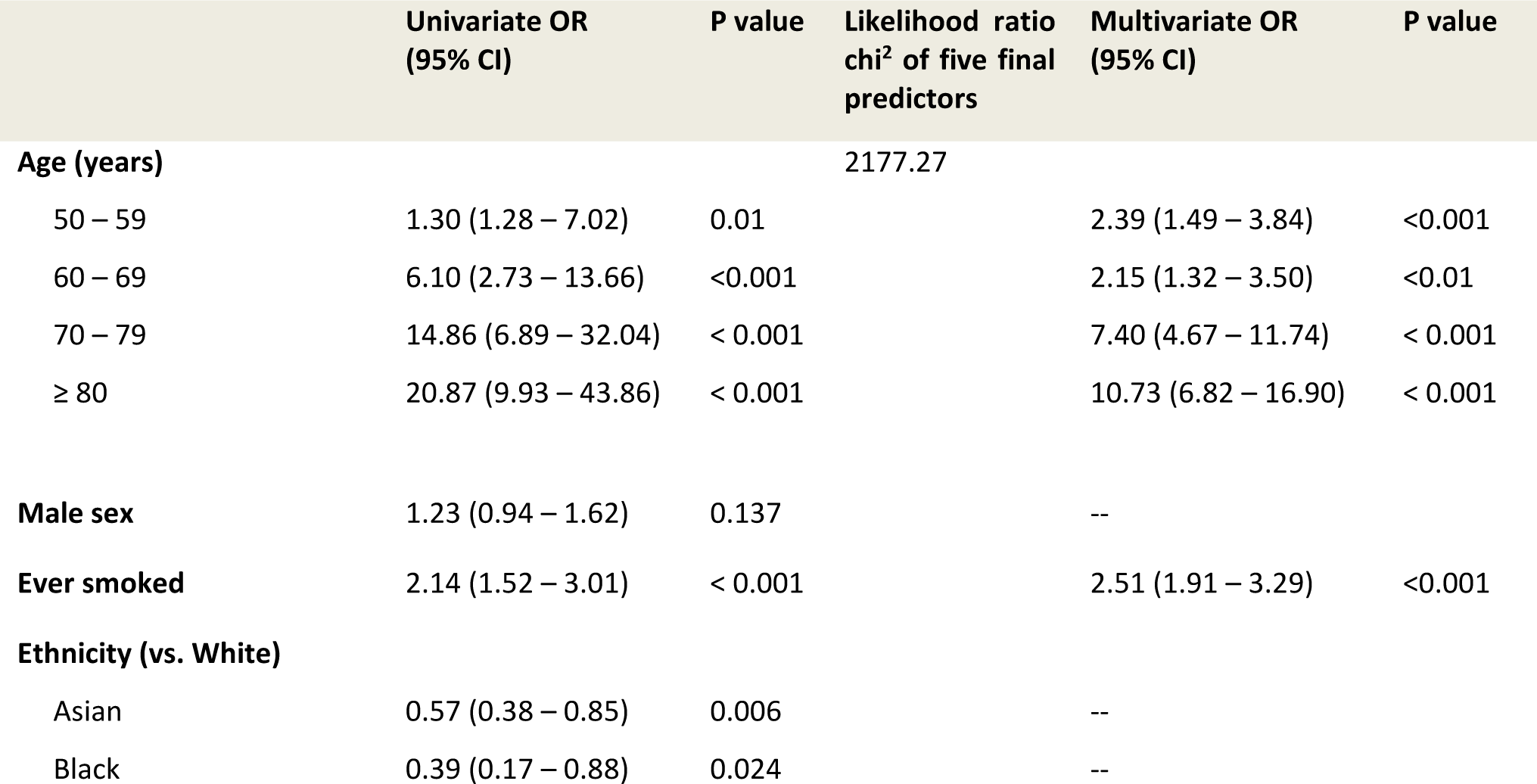
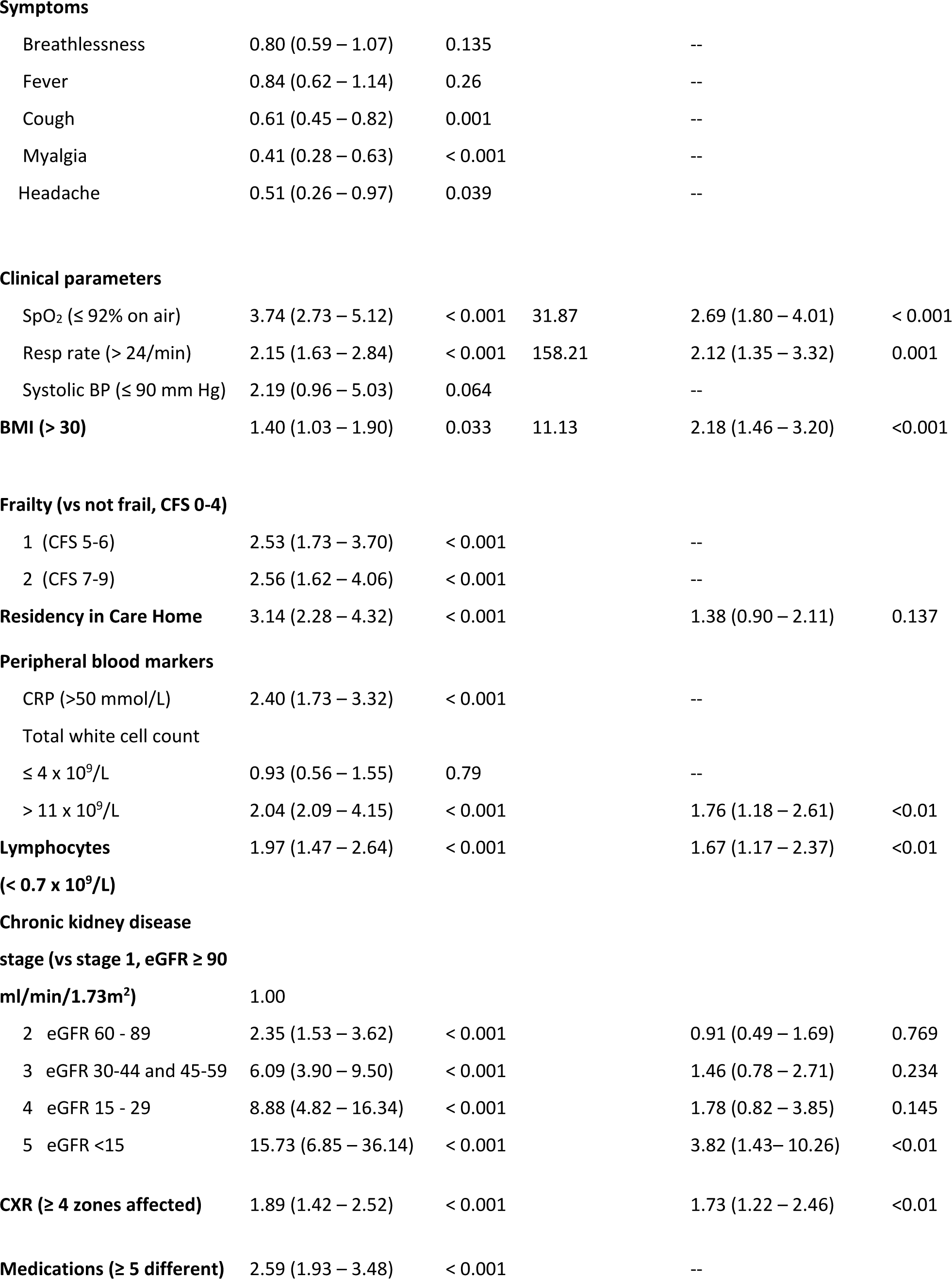
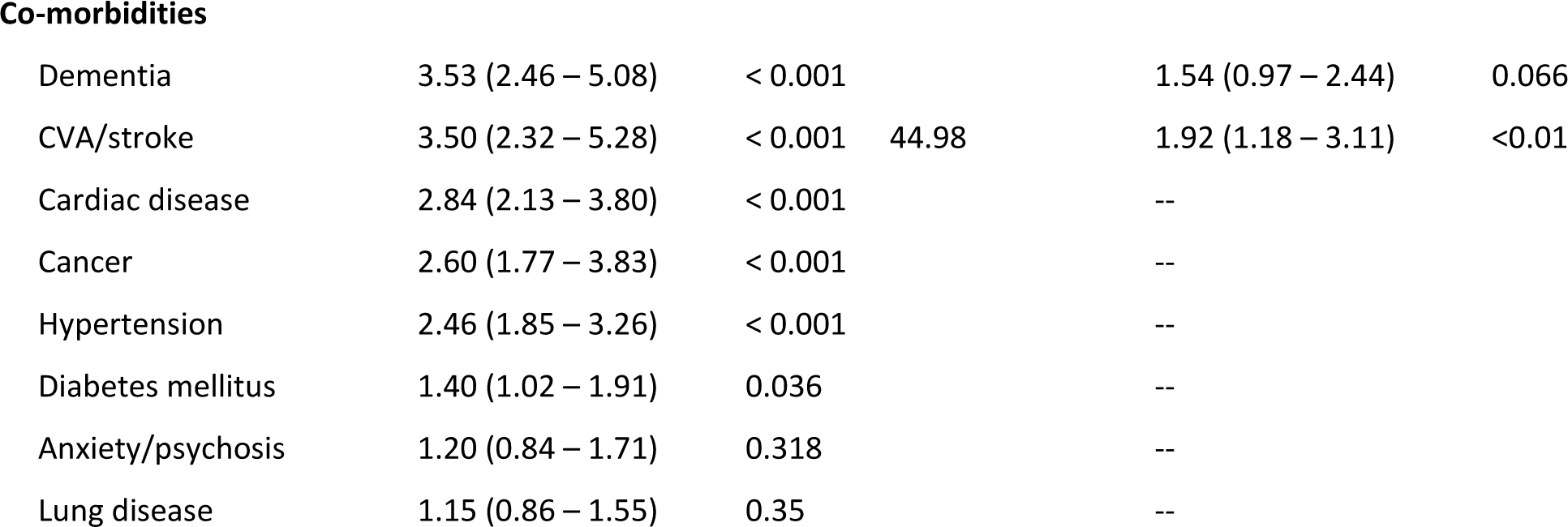
Risk factors of mortality in the derivation cohort (N = 983)

White Caucasian ethnicity constituted 77.3% (760/983) of the whole cohort while Asian, Black and other minor ethnicities (BAME) represented 16.5%, 4.5% and 1.7% respectively. Overall, White ethnicity was associated with the highest proportion of non-survivors (85.0%); in comparison, patients of Asian (OR 0.57, 95% CI: 0.38 – 0.85, *P*<0.01) or Black (OR 0.39, 95% CI: 0.17 – 0.88, *P* <0.05) background in this cohort were not at a higher risk of death from COVID-19. The proportion of non-survivors within each ethnic group was also highest in White (32.9%), followed by Asian (21.6%), Black (15.9%) and other minority groups (17.6%) (*P*<0.01). Of note, White patients were significantly older by median age (74, IQR 58-85) compared to Asian (57, IQR 46-71; *P*<0.0001) or Black (58, IQR 50-72; *P*<0.001).

Care home residency (204/983; 20.8%) was more common amongst non-survivors (*P* <0.001) and was associated with an unadjusted risk of death of 3.14 (95% CI: 2.28 – 4.32, *P*<0.001). Based on frailty data from 644 patients aged 65 or older, the univariate OR of this predictor for death was 2.52 (95% CI: 1.73 – 3.69; *P*<0.001) in those with a group 1 frailty score and 2.56 (95% CI: 1.62 – 4.06; *P*<0.001) in those with a group 2 frailty score (table 1).

The median time from symptom onset to presentation for the derivation cohort was 6 days (IQR 2.0 – 11.0), with no difference between survivors and non-survivors. The four commonest reported symptoms were fever (61%), breathlessness (57.9%), cough (52.8%) and myalgia (21.7%). Tachypnoea (respiratory rate >24/minute) and hypoxia (SpO2 ≤92% on ambient air) were evident in 35.9% and 31.4% of patients respectively, and associated with crude OR for death of 2.15 (95% CI: 1.63 – 2.84, *P*<0.001) and 3.74 (95% CI: 2.73 – 5.12, *P*<0.001) respectively. C-reactive protein >50 mg/L was more frequently documented in non-survivors (76.7% vs. 58.5%; *P*<0.001) and associated with an unadjusted OR for mortality of 2.40 (95% CI: 1.73 – 3.32, *P*<0.001). Lymphopenia was similarly more common in non-survivors (44.7% vs. 29.1%; *P*<0.001). with an unadjusted OR for death of 1.97 (95% CI: 1.47 – 2.64; *P*<0.001). A baseline chest radiograph (CXR) was available in 91% (895/983) patients; abnormalities in ≥4 radiographic zones were evident in 338 (37.8%) of cases and was associated with increased mortality on univariate analysis (OR 1.89, 95% CI 1.42 – 2.52, *P*<0.001).

### Multivariate regression for independent risk factors of mortality

The bootstrapped multivariable regression analysis included the whole derivation cohort of 983 patients comprising 689 (70.1%) survivors and 294 (29.9%) non-survivors with complete or multiply imputed values for data. Older age, CKD stage 5, baseline hypoxia, elevated BMI, tachypnea, leucocytosis and a history of stroke were identified as the strongest independent predictors of mortality (table 1). In particular, age ≥70 had the highest OR for death over the other individual variables. Following adjustment for co-variate effects, Care Home residency was excluded as a predictor of mortality; cut-point analysis also showed that individual blood markers were not independently predictive.

### Iterative modelling to construct an 11-predictor and 5-predictor risk prediction scores

We used values from 770 patients with complete data in the derivation cohort to develop an initial 11-predictor score that ranged from 1 – 18 points (appendix 4). The lowest score of 1 point reflected the KDIGO categorisation of CKD. [11] In this score, the correlation between increasing COVID-19 severity and in-hospital mortality followed a linear dose response relationship, particularly between a score of 3 (below which no deaths occurred) and 12 (above which no patient survived), and an accuracy (AUROC) of 0.84 (figure 1). A shorter 5- predictor score based solely on clinical parameters and scaled from 0 – 8 points was also developed (table 2; figure 1). This short score, abbreviated to **SOARS** (<u>S</u>pO2, <u>O</u>besity, <u>A</u>ge, <u>R</u>espiratory rate, <u>S</u>troke history), demonstrated an AUROC of 0.82 and was retained for further evaluation as a practical pre-hospital risk stratification tool.

**Table 2.** SOARS score (5 predictors; range: 0 – 8 points) for predicting in-hospital COVID-19 death.

| PREDICTOR | POINTS |
| --- | --- |
| **SpO<sub>2</sub>** |  |
| > 92% on air | 0 |
| ≤ 92% on air | 1 |
| **Obesity (BMI >30)** |  |
| absent | 0 |
| present | 1 |
| **Age** |  |
| < 50 | 0 |
| 50 – 59 | 1 |
| 60 – 69 | 2 |
| 70 – 79 | 3 |
| ≥ 80 | 4 |
| **Respiratory rate** |  |
| ≤ 24/min | 0 |
| > 24/min | 1 |
| **Stroke/CVA** |  |
| absent | 0 |
| present | 1 |

**Figure 1.**
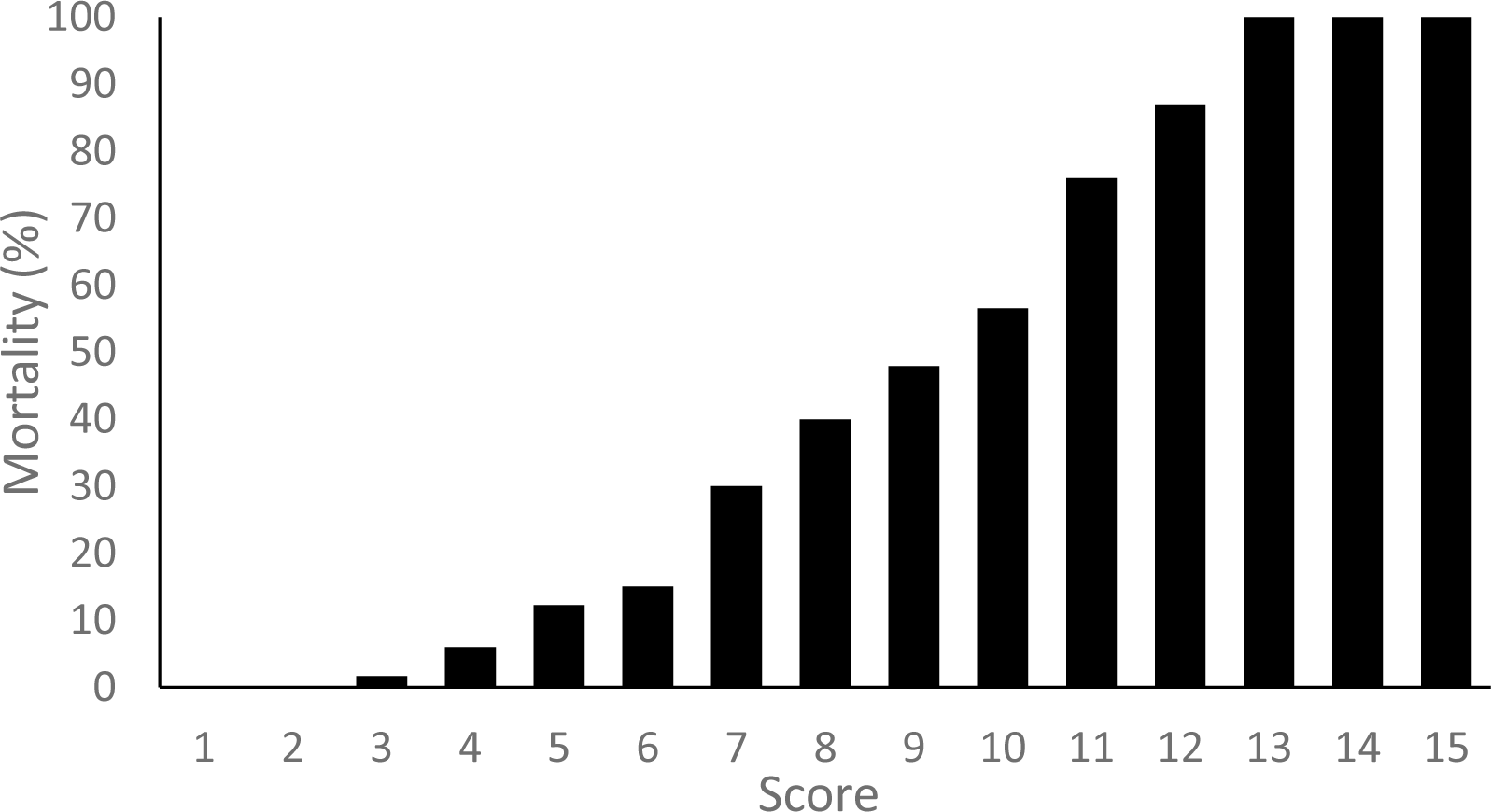
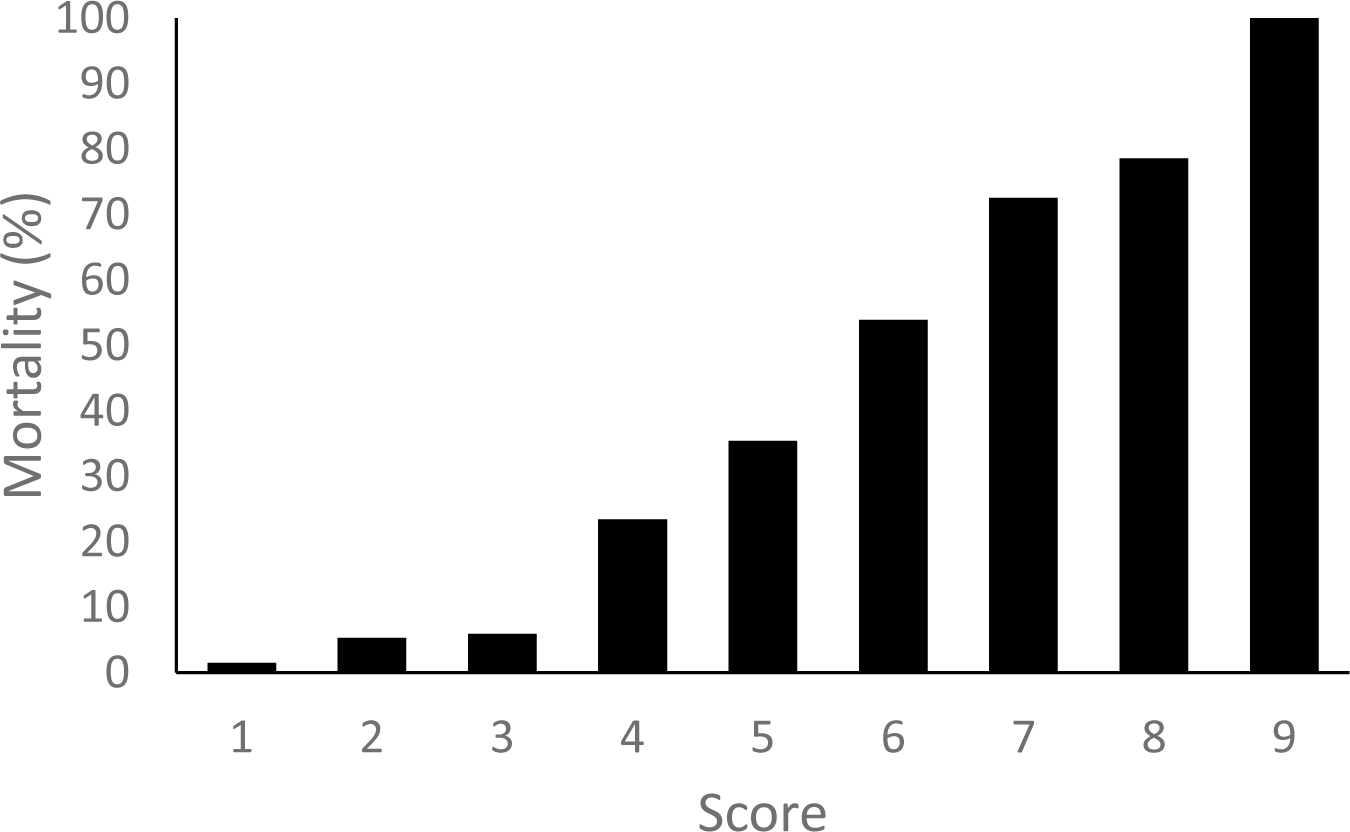
In-hospital mortality in the derivation cohort stratified by the 11-predictor score (AUROC 0.84) In-hospital mortality in the derivation cohort stratified by the 5-predictor SOARS score (AUROC 0.82)

### External validation and performance of the SOARS score

The performance metrics of the long 11-predictor and 5-predictor (SOARS) scores were assessed by their ability to discriminate for in-hospital mortality against both the ISARIC and Aintree validation cohorts (table 3). The longer score showed higher discrimination in the Aintree (AUROC 0.87) than the ISARIC cohort (0.77). The SOARS score had a slightly lower AUROC against both cohorts, namely 0.80 (Aintree) and 0.74 (ISARIC). In comparison, the performance of other scores based solely on different cut-offs of age were associated with inferior discriminatory ability. Comparison of some of the main population parameters between the derivation and both external cohorts are shown in appendix 5.

**Table 3.** Discriminatory performance (area under the receiver operating characteristic; AUROC) of different risk stratification models for predicting COVID-19 in-hospital mortality.

| Cohort | 11-predictor | 5-predictor (SOARS) | Age ≥ 80 | Age 70-79 | Age ≥ 50 |
| --- | --- | --- | --- | --- | --- |
| Derivation | DD0.84 | 0.82 | 0.73 | 0.76 | 0.73 |
| Aintree<br>(validation) | 0.87 | 0.80 | 0.78 | 0.69 | 0.57 |
| ISARIC<br>(validation) | 0.77 | 0.74 | 0.68 | 0.70 | 0.63 |

The mortality rate at each level of the SOARS score in the derivation and both validation cohorts are shown in table 4. For increased applicability, the SOARS score results were further categorised into three risk classes – low (SOARS 0 – 1), moderate (SOARS 2) and high (SOARS ≥3) (table 5). Between 2.3 – 3.2% of patients scoring 0 or 1 (low risk) in each of the three cohorts died due to COVID-19 whereas 2.3 – 6.3% of those scoring 2 (moderate risk) failed to survive to discharge. Overall, 9 out of every 10 deaths in each of the derivation and validation cohorts had a SOARS score of 3 or greater. Within this broad range of increasing scores, the highest proportion of deaths was encountered at SOARS score 4 in both the derivation cohort and ISARIC validation cohort (28.3% and 34.5% respectively) and at SOARS score 5 in the Aintree validation cohort (30.9%).

**Table 4.** Mortality in the derivation and validation cohorts at different levels of SOARS.

| Score | Derivation cohort<br>(N = 821; deaths = 258) |  |  | Aintree validation cohort<br>(N = 290; deaths = 94) |  |  | ISARIC validation cohort<br>(N = 14,231; deaths = 4319) |  |  |
| --- | --- | --- | --- | --- | --- | --- | --- | --- | --- |
|  | No. of deaths<br>(n/N) | Mortality rate (%) | Proportion of deaths (%) | No. of deaths<br>(n/N) | Mortality rate (%) | Proportion of deaths (%) | No. of deaths<br>(n/N) | Mortality rate (%) | Proportion of deaths (%) |
| 0 | 1/69 | 1.4% | 0.4% | 1/12 | 8.3% | 1.1% | 34/833 | 4.1% | 0.8% |
| 1 | 5/94 | 5.3% | 1.9% | 2/23 | 8.7% | 2.1% | 94/1529 | 6.1% | 2.2% |
| 2 | 6/102 | 5.9% | 2.3% | 3/37 | 8.1% | 3.2% | 273/1879 | 14.5% | 6.3% |
| 3 | 29/124 | 23.4% | 11.2% | 11/60 | 18.3% | 11.7% | 650/2577 | 25.2% | 15.1% |
| 4 | 73/206 | 35.4% | 28.3% | 24/72 | 33.3% | 25.5% | 1490/4013 | 37.1% | 34.5% |
| 5 | 63/117 | 53.9% | 24.4% | 29/48 | 60.4% | 30.9% | 1170/2402 | 48.7% | 27.1% |
| 6 | 58/80 | 72.5% | 22.6% | 16/30 | 53.3% | 17.0% | 571/939 | 60.8% | 13.2% |
| 7 | 22/28 | 78.6% | 8.5% | 6/6 | 100.0% | 6.4% | 36/58 | 62.1% | 0.8% |
| 8 | 1/1 | 100.0% | 0.4% | 2/2 | 100.0% | 2.1% | 1/1 | 100.0% | 0.0% |

**Table 5.**
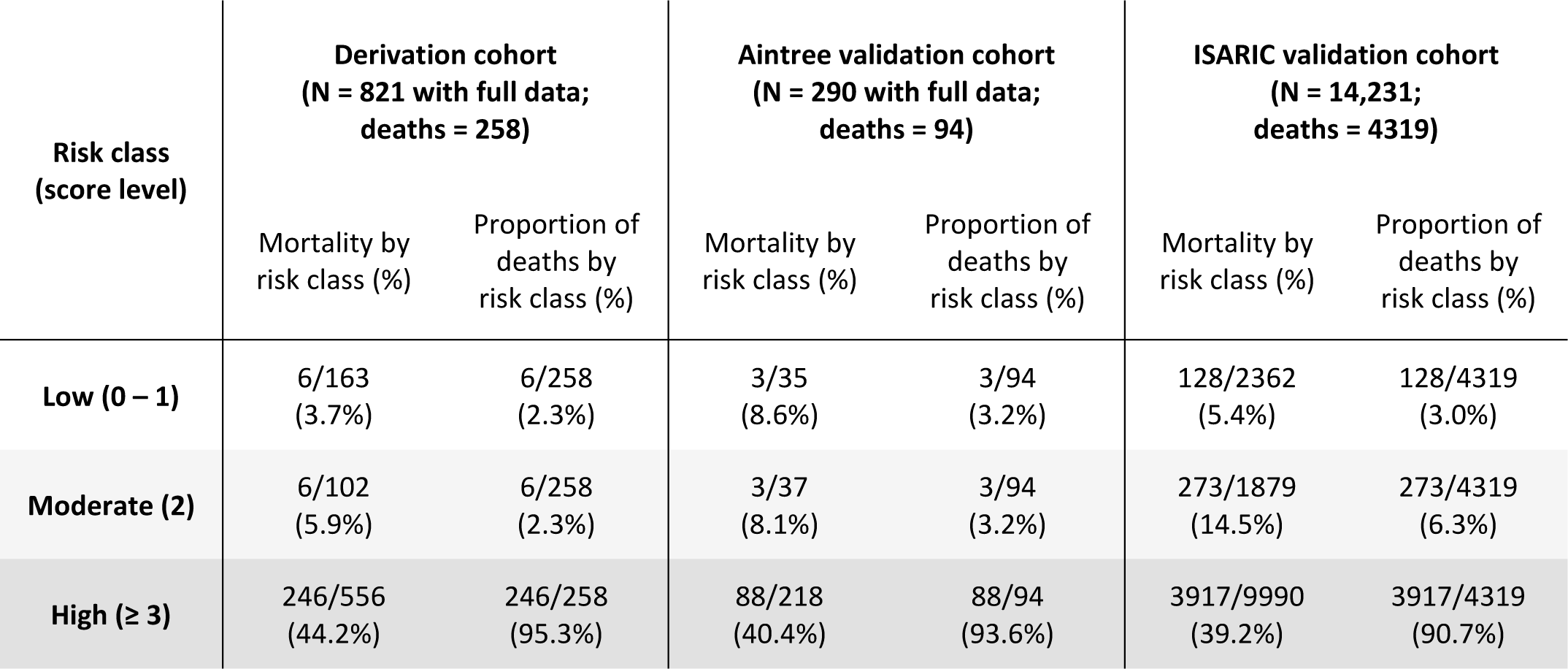
COVID-19 mortality risk stratification based on SOARS score.

Sensitivity thresholds calculated for the larger validation (ISARIC) cohort showed that the low risk class (SOARS 0 – 1) comprised 16.6% of patients with an in-hospital mortality of 5.4%, sensitivity and negative predictive value (NPV) for ruling out death of 99.2% and 95.9% respectively (table 6). By comparison, 29.8% of patients were classified within the moderate risk class; their mortality rate was 14.5%, sensitivity 97.0% and NPV 94.6%. The high risk class comprised 70.2% of the validation cohort who scored across a wide range of SOARS scores, The NPV and specificity for a prediction of death were thus more variable, from 90.5% and 38.7% respectively for a SOARS score of 3, to 69.8% and 99.8% respectively at the other end of the scale when the SOARS score was 7. Only one patient scored 8; they survived to discharge.

**Table 6.** Sensitivity analysis of the SOARS score for predicting mortality in the ISARIC validation cohort.

| Score cut-off | Patients N (% of total) | True pos | True neg | False pos | False neg | Sensitivity | Specificity | PPV | NPV | Cumulative mortality (%) |
| --- | --- | --- | --- | --- | --- | --- | --- | --- | --- | --- |
| >0 | 833 (5.9%) | 4284 | 799 | 9114 | 34 | 99.2% | 8.1% | 32.0% | 95.9% | 34/833 (4.1%) |
| >1 | 2362 (16.6%) | 4190 | 2234 | 7679 | 128 | 97.0% | 22.5% | 35.3% | 94.6% | 128/2362 (5.4%) |
| >2 | 4241 (29.8%) | 3917 | 3840 | 6073 | 401 | 90.7% | 38.7% | 39.2% | 90.5% | 401/4241 (9.5%) |
| >3 | 6818 (47.9%) | 3267 | 5767 | 4146 | 1051 | 75.7% | 58.2% | 44.1% | 84.6% | 1051/6818 (15.4%) |
| >4 | 10831 (76.1%) | 1777 | 8290 | 1623 | 2541 | 41.2% | 83.6% | 52.3% | 76.5% | 2541/10831 (23.5%) |
| >5 | 13233 (93.0%) | 607 | 9522 | 391 | 3711 | 14.1% | 96.1% | 60.8% | 72.0% | 3711/13233 (28.0%) |
| >6 | 14172 (99.6%) | 36 | 9890 | 23 | 4282 | 0.8% | 99.8% | 61.0% | 69.8% | 4282/14172 (30.2%) |
| >7 | 14231 (100.0%) | 0 | 9912 | 1 | 4318 | 0.0% | 100.0% | 0.0% | 69.7% | 4318/14231 (30.3%) |

Reliability of the risk estimates in the ISARIC cohort, modelled as calibration or goodness-of-fit between expected (predicted) and observed outcomes using SOARS, showed a calibration slope of 0.70, calibration-in-the-large (CITL) 0.02 and an expected-to-observed (E:O) ratio of 0.990 (figure 2). Calibration was slightly improved in the Aintree cohort with a slope of 0.80, CiTL −0.16 and E:O ratio of 1.06. A LOWESS (locally weighted scatterplot smoothing algorithm) curve was generated to show differences between these outcomes in both cohorts. The plot characteristics suggested that the model, whilst demonstrating good concordance, had greater predictive accuracy in low to moderate risk patients whose predicted probability of mortality was under 40%. Conversely, overestimation of mortality risk was evident in patients in the high risk group.

**Figure 2.**
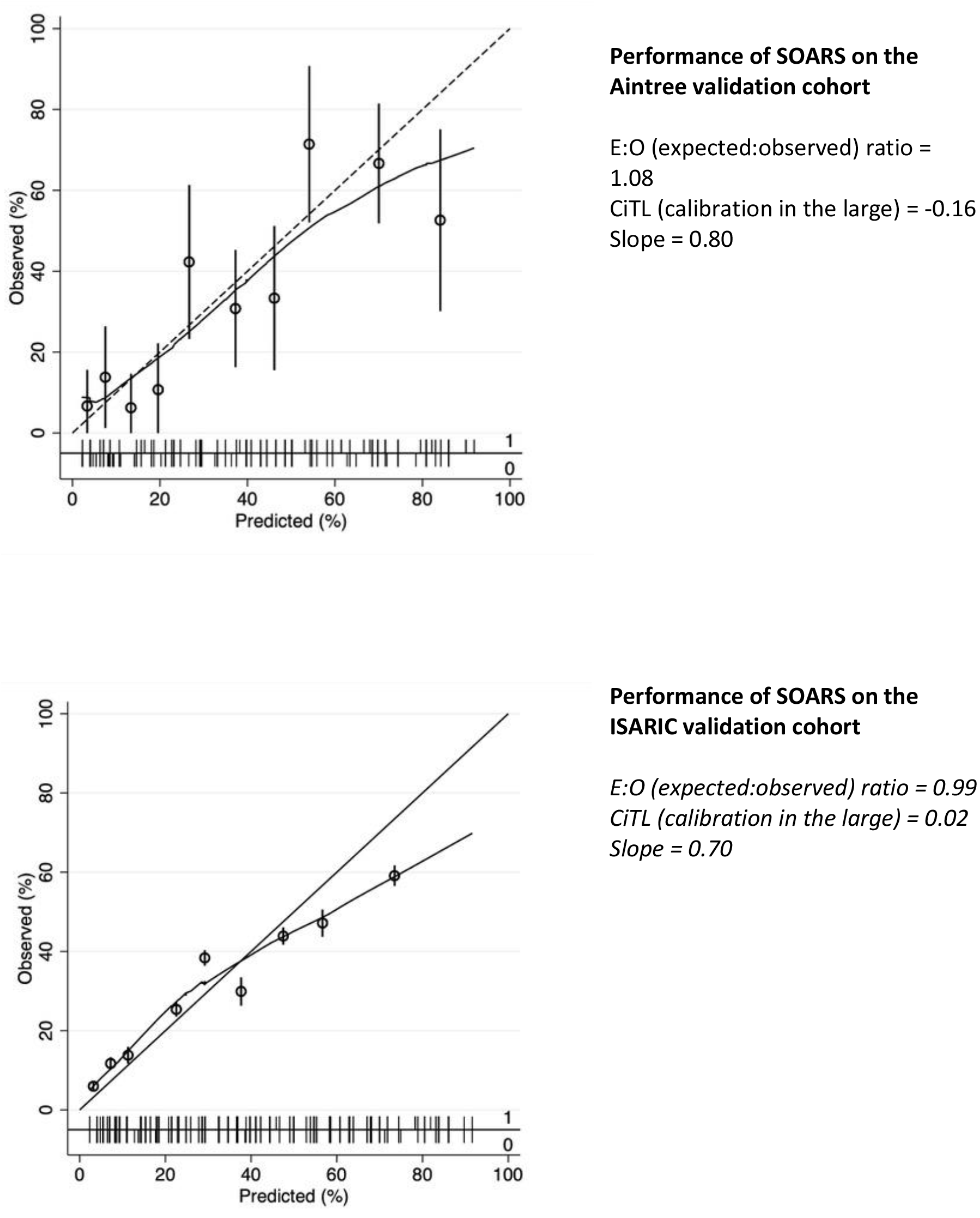
Calibration accuracy of the SOARS score on external validation cohorts Performance of SOARS on the Aintree validation cohort.

## DISCUSSION

We show that prognostic evaluation of a small panel of baseline clinical and demographic characteristics of patients with COVID-19 enables their subsequent risk of in-hospital death to be quantified across three strata of risk. Findings were obtained by applying the 5-predictor SOARS (SpO2, Obesity, Age, Respiratory rate, Stroke history) score to a large random sample of the ISARIC cohort and a smaller single-hospital cohort from Aintree, both with individual-level data. Our objective was to enable risk stratification to be undertaken early, ideally pre-hospitalisation, during the encounter with COVID-19 suspected individuals. This role is not met by currently available prediction tools that rely on laboratory measurements.

The SOARS score discriminated well for COVID-19 mortality and its simplicity obviated the need for complex calculations. It also retained good predictive accuracy in two external validation cohorts, with performance metrics that were primarily reflected in its high negative predictive values for mortality amongst patients with the lowest risk scores (0 or 1). This characteristic is consistent with a high accuracy for predicting a non-fatal outcome in its key target group, namely individuals with milder COVID-19.

Patients who score SOARS 0 or 1 could be discharged home with advice to re-establish urgent contact if their symptoms worsened. Patients stratified as moderate risk (SOARS 2) could be virtually monitored with a pre-defined plan for care escalation if specific thresholds relating to deteriorating symptoms or self-recorded SpO2 were triggered. Patients in the high-risk class (score ≥3) are highly likely to be symptomatic and would, in all probability, be referred directly to the ED for hospital-based management. Thus, the target individuals for the SOARS score are those with a low or moderate risk of COVID-19 mortality.

Our data concur with other reports that advanced age is the strongest predictor of death from COVID-19. [12-15] The sharp increase in mortality in patients within the derivation cohort who were in or beyond their seventh decade of life was reflected in the magnitude of their respective adjusted odds ratios for in-hospital death, namely 7.4 (aged 70 – 79) and 10.7 (aged ≥80). Even so, predictive models generated with age as the lone variable showed poorer discriminatory ability than the SOARS score.

The early development of physiological abnormalities in COVID-19 does not always result in timely clinical presentation. In our study, two measures of acute physiological perturbation proved to be important predictors of COVID-19 mortality: hypoxia and tachypnoea. Although persistent hypoxia is more common in non-survivors of COVID-19, its relationship with tachypnoea remains incompletely understood. [16,17] Fewer than half of patients with COVID-19 who present to hospital with decreased oxygen saturation report experiencing subjective breathlessness. [18-21] One reason for this observation might be so-called ‘silent hypoxia’ where a blunted symptomatic perception of the effects of hypoxaemia is apparent even when low arterial oxygen tension is evident. [22] This phenomenon may be responsible for delays in seeking clinical attention. Such danger could be mitigated by accurate risk assessment including the measurement and tracking of SpO2 in patients who are deemed to not require immediate hospitalisation. The absence of oxygen determination in the CURB-65 score has been cited as limiting its utility in stratifying COVID-19 patients for outpatient management. [23]

The SOARS score was constructed with data from hospitalised patients as the very low adverse event rate amongst non-admitted cases (for example, in the VH pathway) curtailed the development of a prognostic tool. This issue has previously been highlighted in the context of CRB-65 where low event rates in community studies of pneumonia made predictive inferences difficult to conclude. [4] Other scores that have been used in COVID-19 studies have either not been designed for this disease or have relied heavily on laboratory-measured data. [24-27]

Our multivariate regression model was bootstrapped to reduce overfitting but not penalized prior to external validation. In common with other severity scores for COVID-19, we dichotomized several continuous data parameters which may have potentially obscured non-linear effects between predictors and outcome, contributing to the difference in AUROC values between our derivation and validation cohorts and between both validation cohorts. [6, 24, 26] We also used in-hospital mortality as an unambiguous disease-related primary outcome rather than 30 or 60-day mortality. The better performance of the SOARS score in the smaller Aintree validation cohort compared to the much larger ISARIC cohort may have been due to its more homogeneous case-mix. This comparison suggested that, on balance, the simplicity of a pre-hospital risk prediction tool, provided it retained acceptable accuracy, may outweigh any minor diminution of its performance arising from improved practicality.

Other limitations in the study include the occurrence of missing information despite prospective data collection. The use of multiple imputation to estimate missing values for multivariate regression and the availability of nearly 85% of observations for constructing the risk stratification rule helped to mitigate against underestimating their role. The modest sample size of our derivation cohort was dictated by the incident caseload during the pandemic. However, selective sampling of the pandemic timeline was avoided by including all COVID-19 cases from the initial rise to the subsequent decline in new case numbers over the 11-week study period. Finally, reduced score calibration at the high-risk end suggests that SOARS may overestimate the probability of death in the highest risk cases. However, the principal objective of this score was to enhance frontline decision-making in patients with a low predicted risk of mortality at a time when demand for in-patient resources are likely to be high.

In summary, prognostication using the SOARS score can be undertaken concomitantly with SARS-CoV-2 diagnostic testing to inform clinical triaging, including decisions about the placement of the patient for ongoing care. Analysis of the ISARIC validation cohort in this study showed that between 16.6% and 29.8% (those scoring up to SOARS 1 or 2 respectively) could potentially have avoided admission provided a safe alternative to hospitalization was in place. Prospective studies of SOARS implementation will enable the score to be calibrated against other independent cohorts of COVID-19 patients to examine its performance under conditions that may be unique to different localities. Such an opportunity may soon present itself if SARS-CoV-2 transmission continues to increase in the UK and beyond.

## Data Availability

Data sharing
De-identified data can be shared on request after publication of the manuscript. Requests to be forwarded to ORCID ID: 0000-0001-7845-0173

## Acknowledgements

We are grateful to the healthcare teams whose efforts in the clinical field were fundamental to this work. We would like to thank all the patients involved for their vital their contributions. We are very grateful to the ISARIC Coronavirus Clinical Characterisation Consortium (ISARIC- 4C) Investigators in particular, J Kenneth Baillie (lead investigator) and Malcolm G Semple (chief investigator) for providing access to data for the external validation of our model.

## Contributors

RV, AB, MK and RM developed the clinical algorithm and supervised patient management. FC, RV and AD conceived and designed the investigational plan. FC drafted the manuscript with contributions of intellectual content from RV, AD, LGS, PLM and AB. RV, TV, MK, RM, JS, LGS, ET, HM, SM, NM, SA, ML, AO, CP, RK, TJH, RT, SR, MS and JL collected the data at respective sites. AD, FC and RV examined the data and undertook statistical analyses. All authors have seen and approved the final version of the manuscript. RV is guarantor and attests that all named authors and contributors meet authorship criteria and that no others meeting such criteria have been omitted.

## Patient consent for publication

Not required.

## Ethics approval

Ethical approval was provided by Stanmore Research Ethics Committee, London, England (IRAS ID: 283888). The study is registered with the National Health Service Health Research Authority under the reference 20/HRA/2344.

## Pre-specified study protocol

Included in Supplementary Materials.

## Checklist for the appropriate study type

STROBE and TRIPOD checklists are included as supplementary appendices.

## Provenance and peer review statement

Not commissioned, externally peer reviewed. No part of this work has been written by a medical writer or published in printed or electronic form. Some of the findings of this study have been accepted for presentation at the British Thoracic Society Winter Meeting 2020 to be held in February 2021, at which point the abstract will be published in printed format in a supplement of *Thorax*. None of the results of this study have been posted to a preprint server.

## SUPPLEMENTARY MATERIAL

## Appendix 1. Baseline clinical characteristics of the derivation cohort

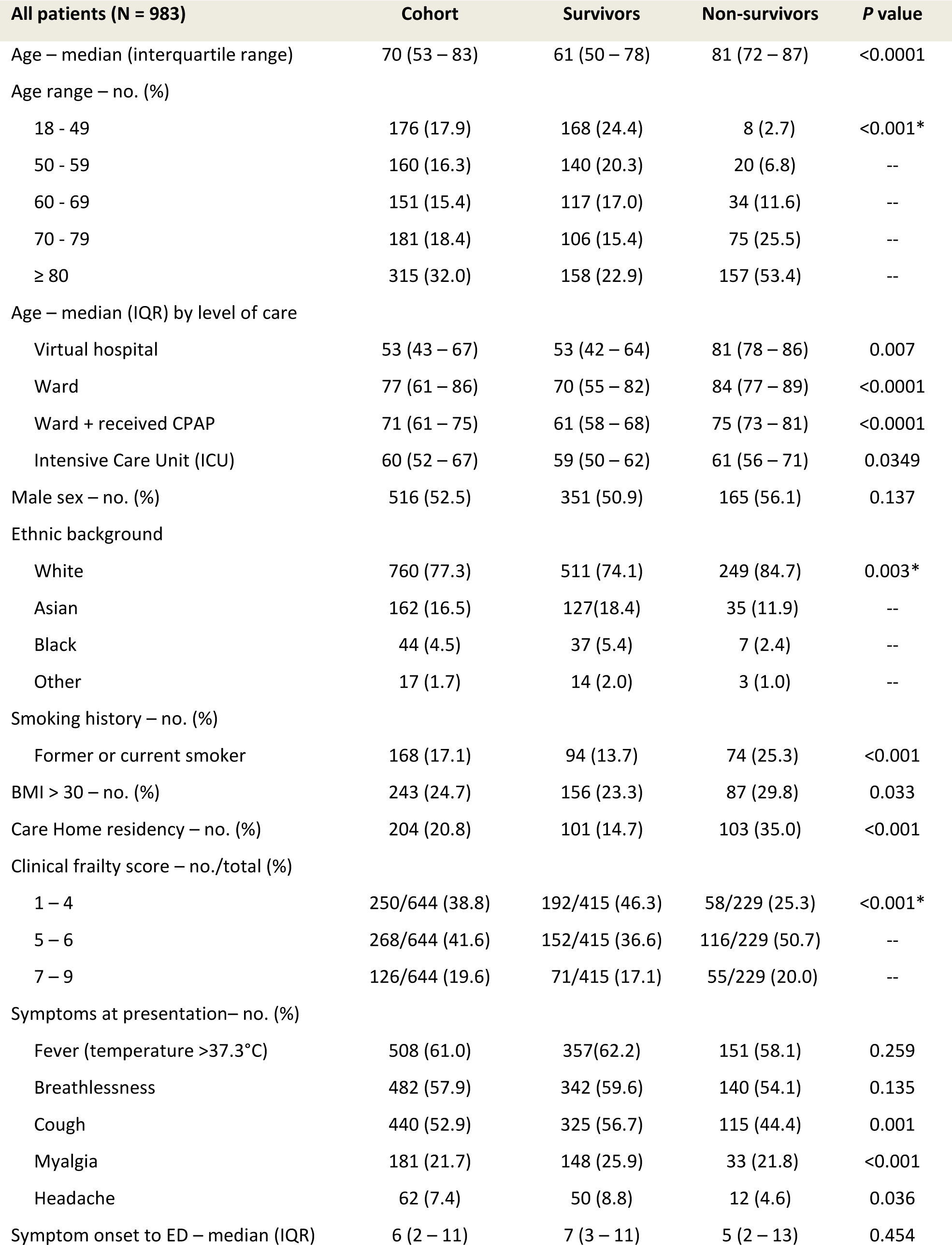
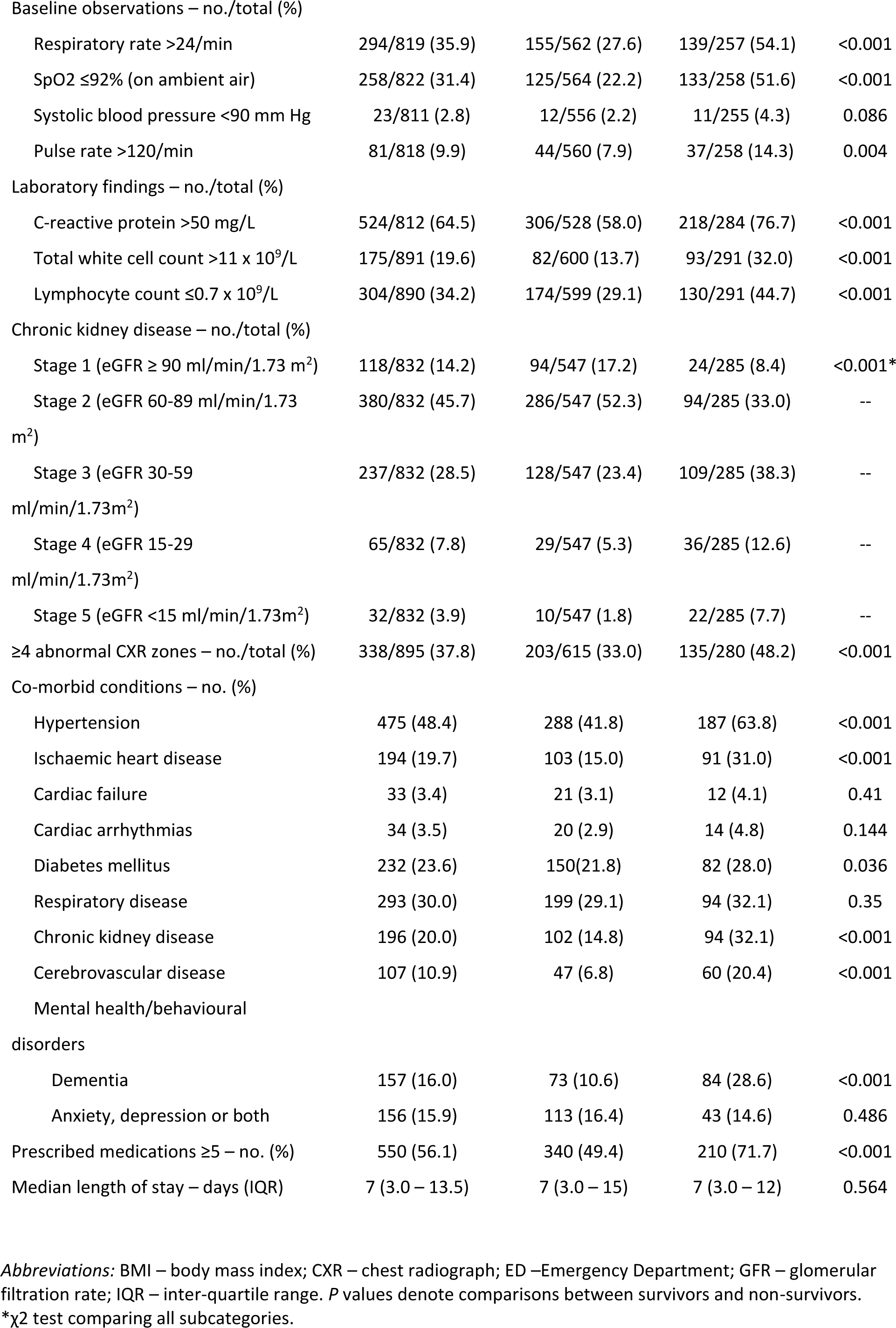

| All patients (N = 983) | Cohort | Survivors | Non-survivors | P value |
| --- | --- | --- | --- | --- |
| Age – median (interquartile range) | 70 (53 – 83) | 61 (50 – 78) | 81 (72 – 87) | <0.0001 |
| Age range – no. (%) |  |  |  |  |
| 18 - 49 | 176 (17.9) | 168 (24.4) | 8 (2.7) | <0.001* |
| 50 - 59 | 160 (16.3) | 140 (20.3) | 20 (6.8) | -- |
| 60 - 69 | 151 (15.4) | 117 (17.0) | 34 (11.6) | -- |
| 70 - 79 | 181 (18.4) | 106 (15.4) | 75 (25.5) | -- |
| ≥ 80 | 315 (32.0) | 158 (22.9) | 157 (53.4) | -- |
| Age – median (IQR) by level of care |  |  |  |  |
| Virtual hospital | 53 (43 – 67) | 53 (42 – 64) | 81 (78 – 86) | 0.007 |
| Ward | 77 (61 – 86) | 70 (55 – 82) | 84 (77 – 89) | <0.0001 |
| Ward + received CPAP | 71 (61 – 75) | 61 (58 – 68) | 75 (73 – 81) | <0.0001 |
| Intensive Care Unit (ICU) | 60 (52 – 67) | 59 (50 – 62) | 61 (56 – 71) | 0.0349 |
| Male sex – no. (%) | 516 (52.5) | 351 (50.9) | 165 (56.1) | 0.137 |
| Ethnic background |  |  |  |  |
| White | 760 (77.3) | 511 (74.1) | 249 (84.7) | 0.003* |
| Asian | 162 (16.5) | 127(18.4) | 35 (11.9) | -- |
| Black | 44 (4.5) | 37 (5.4) | 7 (2.4) | -- |
| Other | 17 (1.7) | 14 (2.0) | 3 (1.0) | -- |
| Smoking history – no. (%) |  |  |  |  |
| Former or current smoker | 168 (17.1) | 94 (13.7) | 74 (25.3) | <0.001 |
| BMI > 30 – no. (%) | 243 (24.7) | 156 (23.3) | 87 (29.8) | 0.033 |
| Care Home residency – no. (%) | 204 (20.8) | 101 (14.7) | 103 (35.0) | <0.001 |
| Clinical frailty score – no./total (%) |  |  |  |  |
| 1 – 4 | 250/644 (38.8) | 192/415 (46.3) | 58/229 (25.3) | <0.001* |
| 5 – 6 | 268/644 (41.6) | 152/415 (36.6) | 116/229 (50.7) | -- |
| 7 – 9 | 126/644 (19.6) | 71/415 (17.1) | 55/229 (20.0) | -- |
| Symptoms at presentation– no. (%) |  |  |  |  |
| Fever (temperature >37.3°C) | 508 (61.0) | 357(62.2) | 151 (58.1) | 0.259 |
| Breathlessness | 482 (57.9) | 342 (59.6) | 140 (54.1) | 0.135 |
| Cough | 440 (52.9) | 325 (56.7) | 115 (44.4) | 0.001 |
| Myalgia | 181 (21.7) | 148 (25.9) | 33 (21.8) | <0.001 |
| Headache | 62 (7.4) | 50 (8.8) | 12 (4.6) | 0.036 |
| Symptom onset to ED – median (IQR) | 6 (2 – 11) | 7 (3 – 11) | 5 (2 – 13) | 0.454 |
| Baseline observations – no./total (%) |  |  |  |  |
| Respiratory rate >24/min | 294/819 (35.9) | 155/562 (27.6) | 139/257 (54.1) | <0.001 |
| SpO<sub>2</sub> ≤92% (on ambient air) | 258/822 (31.4) | 125/564 (22.2) | 133/258 (51.6) | <0.001 |
| Systolic blood pressure <90 mm Hg | 23/811 (2.8) | 12/556 (2.2) | 11/255 (4.3) | 0.086 |
| Pulse rate >120/min | 81/818 (9.9) | 44/560 (7.9) | 37/258 (14.3) | 0.004 |
| Laboratory findings – no./total (%) |  |  |  |  |
| C-reactive protein >50 mg/L | 524/812 (64.5) | 306/528 (58.0) | 218/284 (76.7) | <0.001 |
| Total white cell count >11 x 10<sup>9</sup>/L | 175/891 (19.6) | 82/600 (13.7) | 93/291 (32.0) | <0.001 |
| Lymphocyte count ≤0.7 x 10<sup>9</sup>/L | 304/890 (34.2) | 174/599 (29.1) | 130/291 (44.7) | <0.001 |
| Chronic kidney disease – no./total (%) |  |  |  |  |
| Stage 1 (eGFR ≥ 90 ml/min/1.73 m<sup>2</sup>) | 118/832 (14.2) | 94/547 (17.2) | 24/285 (8.4) | <0.001* |
| Stage 2 (eGFR 60-89 ml/min/1.73 m<sup>2</sup>) | 380/832 (45.7) | 286/547 (52.3) | 94/285 (33.0) | -- |
| Stage 3 (eGFR 30-59 ml/min/1.73m<sup>2</sup>) | 237/832 (28.5) | 128/547 (23.4) | 109/285 (38.3) | -- |
| Stage 4 (eGFR 15-29 ml/min/1.73m<sup>2</sup>) | 65/832 (7.8) | 29/547 (5.3) | 36/285 (12.6) | -- |
| Stage 5 (eGFR <15 ml/min/1.73m<sup>2</sup>) | 32/832 (3.9) | 10/547 (1.8) | 22/285 (7.7) | -- |
| ≥4 abnormal CXR zones – no./total (%) | 338/895 (37.8) | 203/615 (33.0) | 135/280 (48.2) | <0.001 |
| Co-morbid conditions – no. (%) |  |  |  |  |
| Hypertension | 475 (48.4) | 288 (41.8) | 187 (63.8) | <0.001 |
| Ischaemic heart disease | 194 (19.7) | 103 (15.0) | 91 (31.0) | <0.001 |
| Cardiac failure | 33 (3.4) | 21 (3.1) | 12 (4.1) | 0.41 |
| Cardiac arrhythmias | 34 (3.5) | 20 (2.9) | 14 (4.8) | 0.144 |
| Diabetes mellitus | 232 (23.6) | 150(21.8) | 82 (28.0) | 0.036 |
| Respiratory disease | 293 (30.0) | 199 (29.1) | 94 (32.1) | 0.35 |
| Chronic kidney disease | 196 (20.0) | 102 (14.8) | 94 (32.1) | <0.001 |
| Cerebrovascular disease | 107 (10.9) | 47 (6.8) | 60 (20.4) | <0.001 |
| Mental health/behavioural disorders |  |  |  |  |
| Dementia | 157 (16.0) | 73 (10.6) | 84 (28.6) | <0.001 |
| Anxiety, depression or both | 156 (15.9) | 113 (16.4) | 43 (14.6) | 0.486 |
| Prescribed medications ≥5 – no. (%) | 550 (56.1) | 340 (49.4) | 210 (71.7) | <0.001 |
| Median length of stay – days (IQR) | 7 (3.0 – 13.5) | 7 (3.0 – 15) | 7 (3.0 – 12) | 0.564 |

*Abbreviations:* BMI – body mass index; CXR – chest radiograph; ED –Emergency Department; GFR – glomerular filtration rate; IQR – inter-quartile range. *P* values denote comparisons between survivors and non-survivors.
\*χ<sup>2</sup> test comparing all subcategories.

## Appendix 2. In-patient mortality rate by age bracket in the derivation and validation cohorts

| Age group | Derivation cohort | Validation cohorts |  |
| --- | --- | --- | --- |
|  | (N=983) | ISARIC (N=14,231) | Aintree (N=303) |
| < 50 | 4.6% | 5.6% | 9.7% |
| 50 - 59 | 12.5% | 13.8% | 7.9% |
| 60 – 69 | 22.5% | 25.4% | 27.0% |
| 70 – 79 | 41.4% | 37.5% | 44.1% |
| ≥ 80 | 49.8% | 46.2% | 64.9% |
| Overall mortality | 29.9% | 30.9% | 31.7% |

## Appendix 3. In-patient mortality rate by level of care in the derivation cohort (N=983)

|  | Whole cohort | LEVEL OF MAXIMAL CARE |  |  |  |
| --- | --- | --- | --- | --- | --- |
|  |  | VH | Ward | Ward CPAP | ICU |
| N | 983 | 228 | 627 | 41 | 87 |
| Median age (IQR) | 70 (53 – 83) | 53 (43 – 67) | 77 (61 – 86) | 71 (61 – 75) | 60 (52 – 67) |
| Deaths | 294/983<br>(29.9%) | 4/228<br>(1.8%) | 216/627<br>(34.4%) | 20/41<br>(48.8%) | 54/87<br>(62.1%) |
| % of non-survivors by ethnicity at each level of care |  | W 3 (75.0%) | W 194 (90.0%) | W 16 (80.0%) | W 37 (68.5%) |
|  |  | A 1 (25.0%) | A 17 (7.9%) | A 3 (15.0%) | A 13 (24.1%) |
|  |  | B 0 | B 4 | B 0 | B 3 |
|  |  | OTH 0 | OTH 1 | OTH 1 | OTH 1 |

The difference between the median age of patients by level of care, expressed as the Chi-square value with ties, is 150.542 (with 80 degrees of freedom) ( $P < 0.001$ ).
*Abbreviations:* A – Asian; B – Black; CPAP – continuous positive airway pressure; ICU – Intensive Care Unit; IQR – interquartile range; OTH – other ethnicity; VH – virtual hospital; W – White.

## Appendix 4. 11-predictor score (1 – 18 points) for predicting in-hospital COVID-19 death

| **PREDICTOR** | **POINTS** |
| --- | --- |
| **SpO<sub>2</sub>** |  |
| > 92% on air | 0 |
| ≤ 92% on air | 1 |
| **Obesity (BMI >30)** |  |
| absent | 0 |
| present | 1 |
| **Age** |  |
| < 50 | 0 |
| 50 – 59 | 1 |
| 60 – 69 | 2 |
| 70 – 79 | 3 |
| ≥ 80 | 4 |
| **Respiratory rate** |  |
| ≤ 24/min | 0 |
| > 24/min | 1 |
| **Stroke/CVA** |  |
| absent | 0 |
| present | 1 |
| **Ever smoked** |  |
| no | 0 |
| yes | 1 |
| **Dementia** |  |
| no | 0 |
| yes | 1 |
| **CKD stage** |  |
| 1 | 1 |
| 2 | 2 |
| 3 | 3 |
| 4 | 4 |
| 5 | 5 |
| **White cell count >11 x10<sup>9</sup>** |  |
| no | 0 |
| yes | 1 |
| **Lymphocytes ≤ 0.7 x10<sup>9</sup>** |  |
| no | 0 |
| yes | 1 |
| **CXR (≥ 4 zones affected)** |  |
| no | 0 |
| yes | 1 |

## Appendix 5. Comparison of key parameters between the derivation and validation cohorts

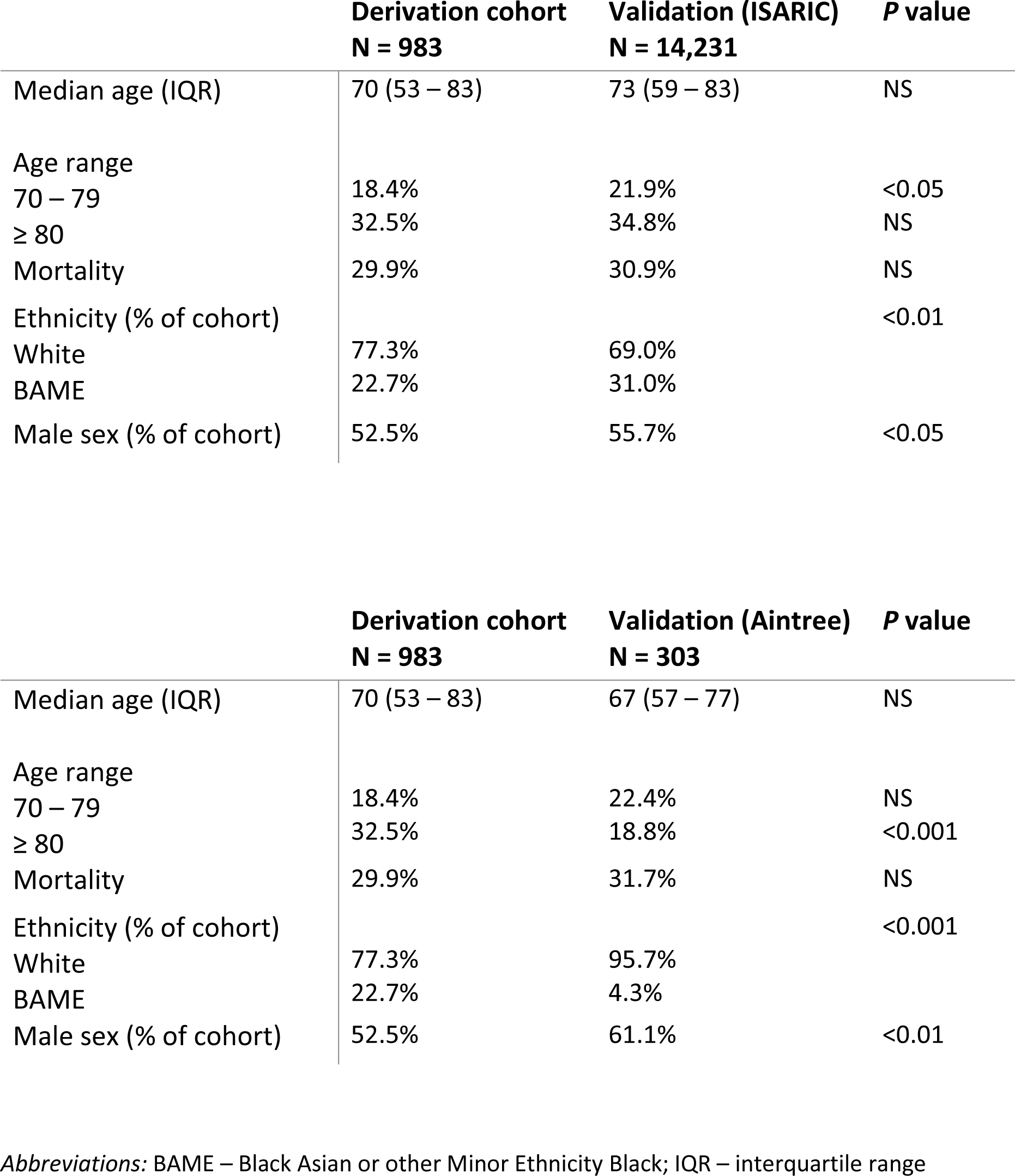

|  | **Derivation cohort**<br>**N = 983** | **Validation (ISARIC)**<br>**N = 14,231** | **P value** |
| --- | --- | --- | --- |
| Median age (IQR) | 70 (53 – 83) | 73 (59 – 83) | NS |
| Age range |  |  |  |
| 70 – 79 | 18.4% | 21.9% | <0.05 |
| ≥ 80 | 32.5% | 34.8% | NS |
| Mortality | 29.9% | 30.9% | NS |
| Ethnicity (% of cohort) |  |  | <0.01 |
| White | 77.3% | 69.0% |  |
| BAME | 22.7% | 31.0% |  |
| Male sex (% of cohort) | 52.5% | 55.7% | <0.05 |

|  | **Derivation cohort**<br>**N = 983** | **Validation (Aintree)**<br>**N = 303** | **P value** |
| --- | --- | --- | --- |
| Median age (IQR) | 70 (53 – 83) | 67 (57 – 77) | NS |
| Age range |  |  |  |
| 70 – 79 | 18.4% | 22.4% | NS |
| ≥ 80 | 32.5% | 18.8% | <0.001 |
| Mortality | 29.9% | 31.7% | NS |
| Ethnicity (% of cohort) |  |  | <0.001 |
| White | 77.3% | 95.7% |  |
| BAME | 22.7% | 4.3% |  |
| Male sex (% of cohort) | 52.5% | 61.1% | <0.01 |

*Abbreviations:* BAME – Black Asian or other Minor Ethnicity Black; IQR – interquartile range

## Appendix. Pre-specified study protocol for clinical data collection

**PREDICT COVID Clinical Data Collection protocol**

**Date: 1**^**st**^ **March 2020**

### Aims

1. To prospectively collect data on all adults (> 18 years) with laboratory-confirmed SARS-CoV-2 infection (COVID-19) presenting to Watford Hospital, West Hertfordshire NHS Trust, during the first wave of the SARS-CoV-2 pandemic, including an outcome of in-hospital death or hospital discharge,
2. To develop a prognostic (risk prediction) score using the above derivation data,
3. To construct a practical clinical scoring system for predicting mortality (and risk of morbidity) after validation of score against external, i.e. independent cohorts of COVID-19 patients from other UK sites,
4. To include and assess outcomes of patients referred to the COVID-19 Virtual Hospital (out-of-hospital monitoring) in the same NHS Trust,
5. To monitor and characterise surviving patients for up to 12 months from the time of confirmation of SARS- CoV-2 infection, including the domains of psychological, physiological and radiological impairment and recovery.

### Primary outcome

In-hospital mortality with minimum 30-day follow-up data.

### Secondary outcomes

Longer term mortality and morbidity: radiological, psychological and cardiorespiratory. This will also be assessed against ongoing health care needs.

### Patient inclusion

All adult patients (aged >18 years) with SARS-CoV-2 real-time reverse transcriptase polymerase chain reaction (rRT-PCR) confirmation.

Completed admission and outcomes at 3 months and 12 months.

### Inclusion criteria

- Readily available patient or clinical characteristic to attending clinicians upon presentation to hospital (Accident & Emergency department, Acute Medical Receiving Unit)
- Blood markers should be commonly measured and results available for review within the first 24 hours of admission
- All parameters relating to oxygen supplementation and advanced respiratory support including one or more of: continuous positive airway pressure (CPAP), bilevel non-invasive ventilation (NIV), high-flow nasal cannula (HFNC) oxygenation and invasive mechanical ventilation (IMV) needs

### Exclusion criteria

All SARS-CoV-2 rRT-PCR negative patients irrespective of clinical suspicion of COVID-19. All individuals aged <18y.

### Selection of candidate variables for initial data collection

Candidate variables were chosen based on knowledge of their potential association/s with SARS-CoV-2 infection and clinical disease (COVID-19). A systematic literature search was undertaken to identify these variables with respect to their predictive association for mortality and other adverse outcomes including COVID-19 severity and disease-related complications such as requirement for critical care and the development of COVID-19-associated acute respiratory distress syndrome (ARDS).

Systematic literature for English language articles in the following search databases: PubMed, EMBASE, WHO Medicus, Web of Science and Google Scholar (particularly for pre-print publications on medRxiv). Search terms included SARS-CoV-2; COVID-19; coronavirus; ARDS; pneumonia; sepsis; influenza; risk prediction; risk score; prognosis; validation. No date restrictions were imposed.

### Statistical analysis for derivation and validation modelling

In analysing the data collected from the derivation (West Herts) cohort, categorical variables will expressed as frequency (%), with significance determined by the Chi-squared test. Continuous variables will be analysed for median (interquartile range) or mean (standard deviation) outcomes and analysed by the t-test, Kruskal-Wallis or Mann-Whitney U test, as appropriate. Missing data in the derivation cohort will be expected even with prospective data collection; missingness of data will be assumed to be at random and handled by multiple imputation by chained equations (MICE) with at least ten imputations, provided the proportion of missingness for the defined parameter constitutes no more than 20% of the cohort. Collated data will be subjected to univariate and multivariate logistic regression in order to determine odds ratios (OR) for in-hospital mortality. The latter will also be internally validated by bootstrapping using a minimum of 1000 re-samples. Predictor interactions will be analysed by appropriate methodology such as the likelihood ratio (LR) test comparing broad and narrow (constrained) models.

All performance metrics against external validation cohorts will be analysed using the prediction score and not with the multivariate regression model. The performance of the derivation model will be assessed for discriminatory ability (area under the receiver operating characteristic, AUROC) and calibration (graphical representation of Hosmer-Lemeshow analysis).

All statistical analyses including risk modelling calculations will be performed using STATA, version 16 (Stata Corp., Texas, USA).

*R Vancheeswaran, F Chua, A Draper, T Vaghela and A Barlow (study design and responsible persons for data analysis and interpretation)*

*MARCH 2020*

#### STROBE Statement—Checklist of items that should be included in reports of *cohort studies*

|  | Item No | Recommendation |
| --- | --- | --- |
| **Title and abstract** | 1 | (a) Indicate the study's design with a commonly used term in the title or the abstract<br>(b) Provide in the abstract an informative and balanced summary of what was done and what was found |
| **Introduction** |  |  |
| Background/rationale | 2 | Explain the scientific background and rationale for the investigation being reported |
| Objectives | 3 | State specific objectives, including any prespecified hypotheses |
| **Methods** |  |  |
| Study design | 4 | Present key elements of study design early in the paper |
| Setting | 5 | Describe the setting, locations, and relevant dates, including periods of recruitment, exposure, follow-up, and data collection |
| Participants | 6 | (a) Give the eligibility criteria, and the sources and methods of selection of participants. Describe methods of follow-up<br>(b) For matched studies, give matching criteria and number of exposed and unexposed |
| Variables | 7 | Clearly define all outcomes, exposures, predictors, potential confounders, and effect modifiers. Give diagnostic criteria, if applicable |
| Data sources/measurement | 8* | For each variable of interest, give sources of data and details of methods of assessment (measurement). Describe comparability of assessment methods if there is more than one group |
| Bias | 9 | Describe any efforts to address potential sources of bias |
| Study size | 10 | Explain how the study size was arrived at |
| Quantitative variables | 11 | Explain how quantitative variables were handled in the analyses. If applicable, describe which groupings were chosen and why |
| Statistical methods | 12 | (a) Describe all statistical methods, including those used to control for confounding<br>(b) Describe any methods used to examine subgroups and interactions<br>(c) Explain how missing data were addressed<br>(d) If applicable, explain how loss to follow-up was addressed<br>(e) Describe any sensitivity analyses |
| **Results** |  |  |
| Participants | 13* | (a) Report numbers of individuals at each stage of study—eg numbers potentially eligible, examined for eligibility, confirmed eligible, included in the study, completing follow-up, and analysed<br>(b) Give reasons for non-participation at each stage<br>(c) Consider use of a flow diagram |
| Descriptive data | 14* | (a) Give characteristics of study participants (eg demographic, clinical, social) and information on exposures and potential confounders<br>(b) Indicate number of participants with missing data for each variable of interest<br>(c) Summarise follow-up time (eg, average and total amount) |
| Outcome data | 15* | Report numbers of outcome events or summary measures over time |
| Main results | 16 | (a) Give unadjusted estimates and, if applicable, confounder-adjusted estimates and their precision (eg, 95% confidence interval). Make clear which confounders were adjusted for and why they were included<br>(b) Report category boundaries when continuous variables were categorized<br>(c) If relevant, consider translating estimates of relative risk into absolute risk for a meaningful time period |
| Other analyses | 17 | Report other analyses done—eg analyses of subgroups and interactions, and sensitivity analyses |
| **Discussion** |  |  |
| Key results | 18 | Summarise key results with reference to study objectives |
| Limitations | 19 | Discuss limitations of the study, taking into account sources of potential bias or imprecision. Discuss both direction and magnitude of any potential bias |
| Interpretation | 20 | Give a cautious overall interpretation of results considering objectives, limitations, multiplicity of analyses, results from similar studies, and other relevant evidence |
| Generalisability | 21 | Discuss the generalisability (external validity) of the study results |
| **Other information** |  |  |
| Funding | 22 | Give the source of funding and the role of the funders for the present study and, if applicable, for the original study on which the present article is based |

\*Give information separately for exposed and unexposed groups.
**Note:** An Explanation and Elaboration article discusses each checklist item and gives methodological background and published examples of transparent reporting. The STROBE checklist is best used in conjunction with this article (freely available on the Web sites of PLoS Medicine at http://www.plosmedicine.org/, Annals of Internal Medicine at http://www.annals.org/, and Epidemiology at http://www.epidem.com/). Information on the STROBE Initiative is available at http://www.strobe-statement.org.

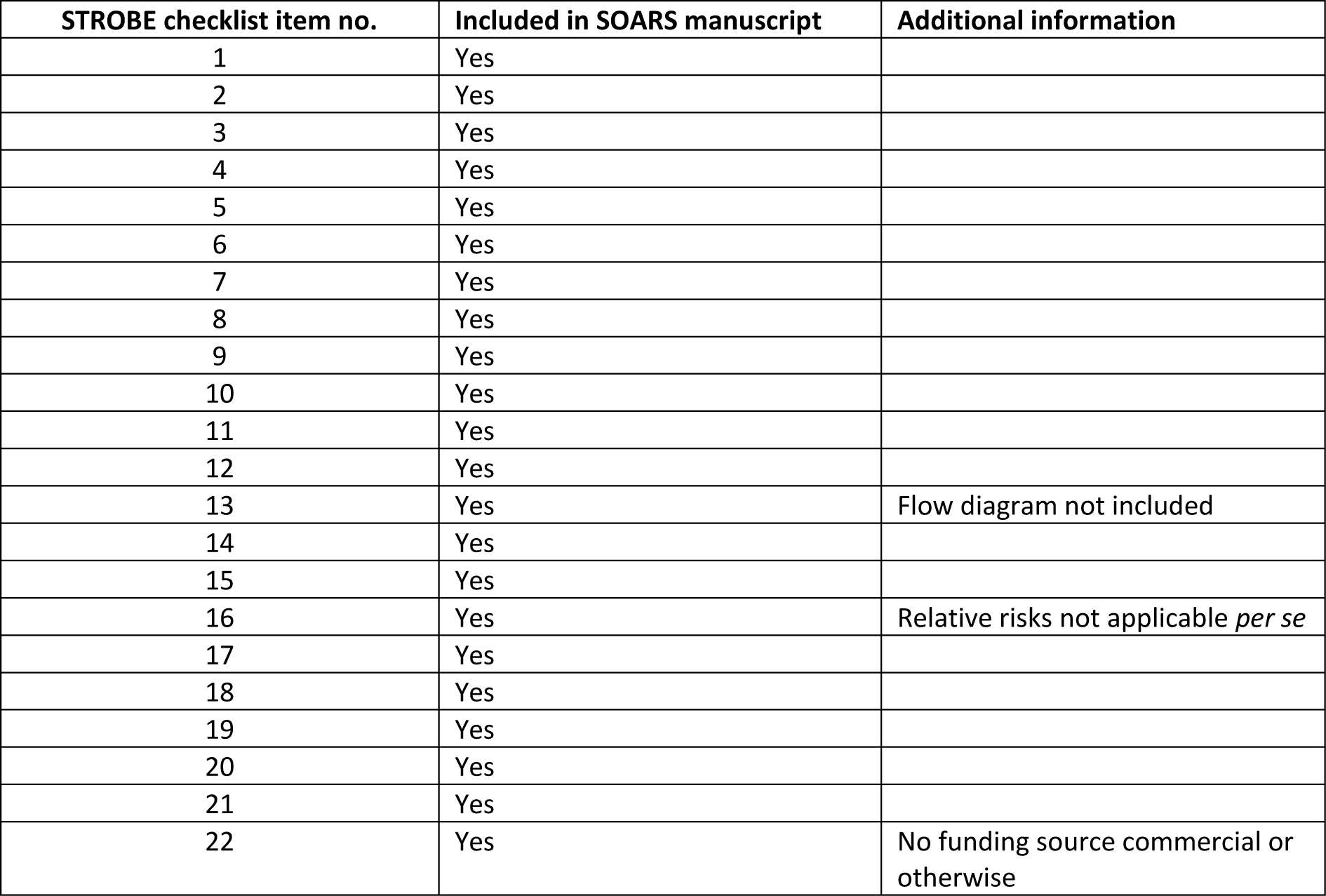

| STROBE checklist item no. | Included in SOARS manuscript | Additional information |
| --- | --- | --- |
| 1 | Yes |  |
| 2 | Yes |  |
| 3 | Yes |  |
| 4 | Yes |  |
| 5 | Yes |  |
| 6 | Yes |  |
| 7 | Yes |  |
| 8 | Yes |  |
| 9 | Yes |  |
| 10 | Yes |  |
| 11 | Yes |  |
| 12 | Yes |  |
| 13 | Yes | Flow diagram not included |
| 14 | Yes |  |
| 15 | Yes |  |
| 16 | Yes | Relative risks not applicable *per se* |
| 17 | Yes |  |
| 18 | Yes |  |
| 19 | Yes |  |
| 20 | Yes |  |
| 21 | Yes |  |
| 22 | Yes | No funding source commercial or otherwise |

#### TRIPOD Checklist: Prediction Model Development and Validation

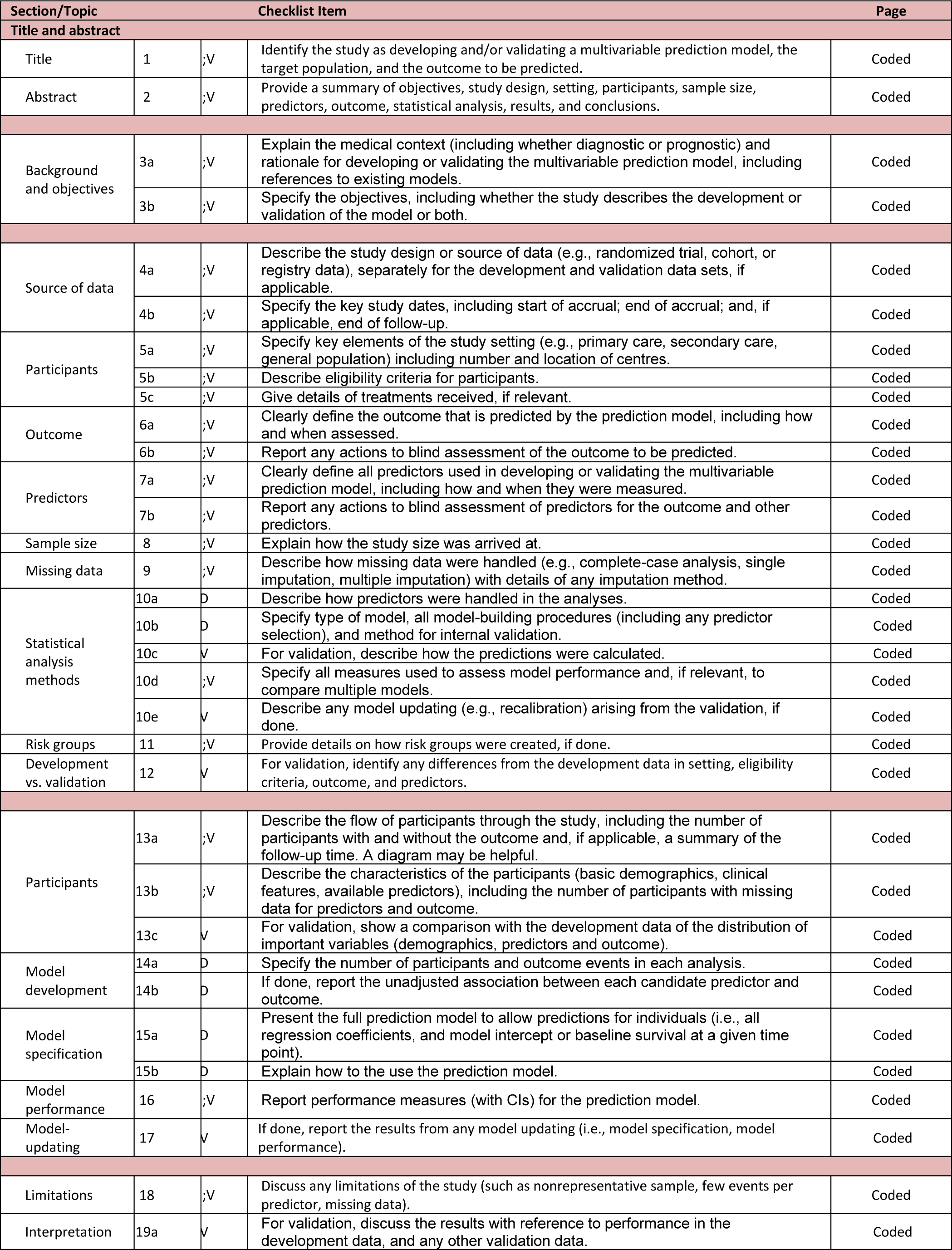
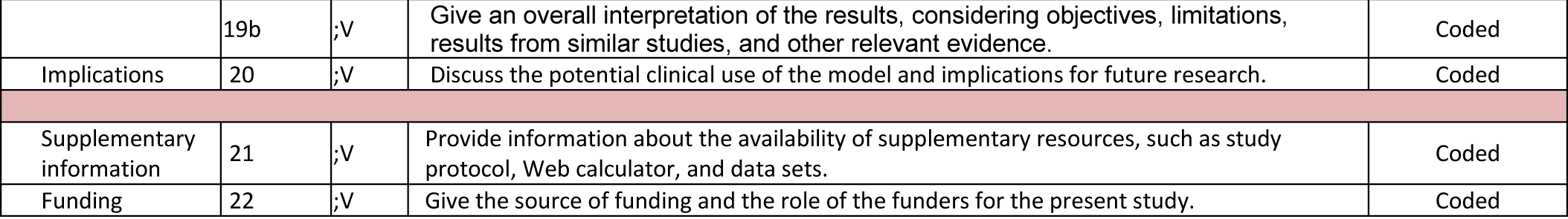

END

## REFERENCES

1. Challen K, Goodacre SW, Wilson A, et al. Evaluation of triage methods used to select patients with suspected pandemic influenza for hospital admission. Emerg Med J 2012; 29: 383–8. doi: 10.3310/hta14460-03

2. Ho PL, Chau PH, Yip PSF, et al. A clinical prediction rule for clinical diagnosis of severe acute respiratory syndrome. Eur Resp J 200; 26: 474–9. doi: 10.1183/09031936.05.1076704

3. Wynants L, Van Calster B, Collins GS, et al. Prediction models for diagnosis and prognosis of covid-19: systematic review and critical appraisal. BMJ 2020; 369: m1328. doi: 10.1136/bmj.m1328

4. McNally M, Curtain J, O’Brien KK, et al. Validity of British Thoracic Society guidance (the CRB-65 rule) for predicting the severity of pneumonia in general practice: systematic review and meta-analysis. Br J Gen Pract 2010; 60: 423–33. doi: 10.3399/bjgp10X532422

5. Lim WS, van der Eerden MM, Laing R, et al. Defining community acquired pneumonia on presentation to hospital: an international derivation and validation study. Thorax 2003; 58: 377–82.

6. Knight S, Ho A, Pius R, et al. Risk stratification of patients admitted to hospital with covid-19 using the ISARIC WHO Clinical Characterisation Protocol: development and validation of the 4C Mortality Score. BMJ 2020; 370: m3339. doi: 10.1136/bmj.m3339

7. National Early Warning Score (NEWS)2. Royal College of Physicians, London. https://www.rcplondon.ac.uk/projects/outputs/national-early-warning-score-news-2. (accessed May 30, 2020).

8. Collins GS, Reitsma JB, Altan DG, et al. Transparent reporting of a multivariable prediction model for individual prognosis or diagnosis (TRIPOD): The TRIPOD statement. Ann Intern Med 2015; 162: 55–63. doi.org/10.1186/s12916-014-0241-z

9. Resche-Rigon M, White IR. Multiple imputation by chained equations for systemically and sporadically missing multilevel data. Stat Methods Med Res 2018; 27: 1634–49. doi: 10.1177/0962280216666564

10. Shipe ME, Deppen SA, Farjah F, et al. Developing prediction models for clinical use using logistic regression: an overview. J Thorac Dis 2019; 11(Suppl 4): S574–84. doi: 10.21037/jtd.2019.01.25

11. Kidney Disease, Improving Clinical Outcomes. KDIGO 2012 Clinical Practice Guideline for the Evaluation and Management of Chronic Kidney Disease. https://kdigo.org/wp-content/uploads/2017/02/KDIGO_2012_CKD_GL.pdf (accessed 1 June 2020).

12. Williamson EJ, Walker AJ, Bhaskaran K, et al. OpenSAFELY: factors associated with COVID-19 death in 17 million patients. Nature 2020; 584: 430–4. doi: 10.1038/s41586-020-2521-4

13. Mikami T, Miyashita H, Yamada T, et al. Risk factors for mortality in patients with COVID-19 in New York City. J Gen Intern Med 2020. doi: 10.1007/s11606-020-05983-z

14. Feng Y, Ling Y, Bai T, et al. COVID-19 with different severities: a multicenter study of clinical features. Am J Respir Crit Care Med 2020; 201: 1380–88. doi: 10.1164/rccm.202002-0445OC

15. Onder G, Rezza G, Brusaferro S. Case-fatality rate and characteristics of patients dying in relation to COVID-19 in Italy. JAMA. 2020; 323: 1775–76. doi:10.1001/jama.2020.4683

16. Xie J, Covassin N, Fan Z, et al. Association between hypoxaemia and mortality in patients with COVID-19. Mayo Clin Proc 2020; 95: 1138–47. doi: 10.1016/j.mayocp.2020.04.006

17. Wu C, Chen X, Cai Y, et al. Risk factors associated with acute respiratory distress syndrome and death in patients with coronavirus disease 2019 pneumonia in Wuhan, China. JAMA Intern Med 2020; 180: 934–43. doi:10.1001/jamainternalmed.2020.0994

18. Richardson S, Hirsch JS, Narasimhan M, et al. Presenting characteristics, comorbidities, and outcomes among 5700 pateints hospitalized with COVID-19 in the New York City area. JAMA 2020; doi:10.1001/jama.2020.6775

19. Guan W, Ni Z, Hu Y, et al. for the China Medical Treatment Expert Group for Covid-19. Clinical characteristics of coronavirus disease in China. N Engl J Med 2020; 382: 1708–20. doi: 10.1056/NEJMoa2002032

20. Zhou F, Yu T, Du R, et al. Clinical course and risk factors for mortality of adult inpatients with COVID-19 in Wuhan, China: a retrospective cohort study. Lancet 2020; 395: 1054–62. doi: 10.1016/S0140-6736(20)30566-3

21. Huang C, Wang Y, Li X, et al. Clinical features of patients infected with 2019 novel coronavirus in Wuhan, China. Lancet 2020. doi: 10.1016/S0140-6736(20)30183-5

22. Tobin MJ, Laghi F, Jubran A. Why COVID-19 hypoxaemia is baffling to physicians. Am J Respir Crit Care Med 2020; 202: 356–60. doi: 10.1164/rccm.202006-2157CP

23. Nguyen Y, Corre F, Honsel V, et al. Applicability of the CURB-65 pneumonia severity score for outpatient treatment of COVID-19. J Infect 2020. doi: 10.1016/j.jinf.2020.05.049

24. Fan G, Tu C, Zhou F, et al. Comparison of severity scores for COVID-19 patients with pneumonia: a retrospective study. Eur Respir J 2020; 56: 2002113. doi: 10.1183/13993003.02113-2020

25. Liang W, Liang H, Ou L, et al. Development and validation of a clinical risk score to predict the occurrence of critical illness in hospitalized patients with COVID-19. JAMA Intern Med. doi:10.1001/jamainternmed.2020.2033

26. Satici C, Demirkol MA, Altunok ES, et al. Performance of pneumonia severity index and CURB-65 in predicting 30-day mortality in patients with COVID-19. Int J Infect Dis 2020; 98: 84–9. doi: 10.1016/j.ijid.2020.06.038

27. Liu S, Yao N, Qiu Y, et al. Predictive performance of SOFA and qSOFA for in-hospital mortality in severe novel coronavirus disease. Am J Emerg Med 2020. doi: 10.1016/j.ajem.2020.07.019

